# A single intramuscular injection of monoclonal antibody MAD0004J08 induces in healthy adults SARS-CoV-2 neutralising antibody titres exceeding those induced by infection and vaccination

**DOI:** 10.1101/2021.08.03.21261441

**Authors:** Simone Lanini, Stefano Milleri, Emanuele Andreano, Sarah Nosari, Ida Paciello, Giulia Piccini, Alessandra Gentili, Adhuna Phogat, Inesa Hyseni, Margherita Leonardi, Alessandro Torelli, Emanuele Montomoli, Andrea Paolini, Andrea Frosini, Andrea Antinori, Emanuele Nicastri, Enrico Girardi, Maria Maddalena Plazzi, Giuseppe Ippolito, Francesco Vaia, Giovanni Della Cioppa, Rino Rappuoli

## Abstract

**Background:** The emerging threat represented by SARS-CoV-2 variants, demands the development of therapies for better clinical management of COVID-19. MAD0004J08 is an extremely potent Fc-engineered monoclonal antibody (mAb) able to neutralise *in vitro* all current SARS-CoV-2 variants of concern (VoCs). This ongoing study, evaluates safety, pharmacokinetics and SARS-CoV-2 sera neutralization effect of MAD0004J08 when administered as single dose intramuscularly in healthy adults.

**Method:** We conducted a dose escalation study with sequential enrolment of three cohorts, each with an increasing dose level of MAD0004J08 (48mg, 100mg and 400mg). Within each cohort, 10 young healthy adults were randomized with 4:1 ratio to a single intramuscular (*i.m.*) injection of MAD0004J08 or placebo. The primary endpoint is the proportion of subjects with severe and/or serious treatment emergent adverse events (TEAEs) within 7 days post-treatment. Secondary endpoints reported in this paper are the proportion of subjects with solicited TEAEs up 7 days post dosing, MAD0004J08 serum concentrations and neutralising activity versus the original SARS-COV-2 Wuhan virus at different timepoints post-dosing. As post-hoc analyses, we compared the sera neutralising titres of subjects who received MAD0004J08 with those of people that had received the COVID-19 BNT162b2 mRNA vaccine in the previous sixty days (n=10) and COVID-19 convalescent patients (n=20), and assessed serum neutralisation activity against the B.1.1.7 (alpha), B.1.351 (beta) and B.1.1.248 (gamma) SARS-CoV-2 variants of concern.

**Findings:** A total of 30 subjects, 10 per cohort, were enrolled and randomized. Data up to 30 days were available and analysed in this report. No severe TEAEs were reported in any of the cohorts in the 7 days post-treatment. MAD0004J08 was detected in the sera of treated subjects within few hours post-administration and reached almost maximal levels on day 8. The geometric mean neutralising titres (GMT) assessed against the original Wuhan virus peaked on day 8 and ranged 226 – 905, 905 – 2,560, and 1,280 – 5,120 for cohort 1, 2 and 3 respectively. The sera neutralising GMT in MAD0004J08 treated subjects in all the three cohorts were found to be 1·5-54·5-fold higher compared to sera from convalescent patients and 1·83– 76·4-fold higher compared to sera from COVID-19 vaccinees. Finally, GMT in MAD0004J08 treated subjects showed high neutralising titres versus the B.1.1.7 (alpha), B.1.351 (beta) and B.1.1.248 (gamma) SARS-CoV-2 VoCs.

**Interpretation:** A single dose administration of MAD0004J08 via *i.m.* route is safe and well tolerated and results in a rapid systemic distribution of the MAD0004J08 and sera neutralising titres higher than COVID-19 convalescent and vaccinated subjects. A single dose administration of MAD0004J08 is also sufficient to effectively neutralise major SARS-CoV-2 variants of concern. Based on these results, a Phase 2-3 trial is ongoing to further assess the safety, dosage, and efficacy of MAD0004J08 in asymptomatic or mild-moderate symptomatic COVID-19 patients.

**Funding:** EU Malaria Fund, Ministero dello Sviluppo Economico, Ministero della Salute, Regione Toscana, Toscana Life Sciences Sviluppo and European Research Council.

**Research in context:** *Evidence before this study:* We searched PUBMED, MEDLINE and MedRxiv for clinical trials, meta-analyses and randomized controlled trials evaluating the antibody neutralization titres vs. different SARS-CoV-2 variants of concern obtained from subjects who received monoclonal antibodies for the treatment of COVID-19 using the following search terms: (“COVID-19” OR “SARS-CoV-2”) AND (“monoclonal antibody” OR “neutralising antibody”) AND (“variants” OR “variants of concern”). No relevant studies were identified.

*Added value of this study:* This is the first human study assessing safety, PK and neutralising potential of MAD0004J08, a monoclonal antibody against SARS-CoV-2 wild type Wuhan virus and variants of concern, administered intramuscularly at low dosages (48, 100 and 400 mg). MAD0004J08 showed to be safe and well tolerated in the tested dose range. Anti-spike antibodies were detected in the sera of tested SARS-CoV-2 negative healthy adults few hours post-injection. In addition, the sera obtained from MAD0004J08treated subjects, showed to have high neutralisation titres against the Wuhan virus, the B.1.1.7 (alpha), B.1.351 (beta) and B.1.1.248 (gamma) variants of concern.

*Implications of all the available evidence:* A potent monoclonal antibody such as MAD0004J08, capable of neutralising multiple variants of concern of SARS-CoV-2 rapidly and long lastingly when given as a single intramuscular injection. The antibody, presently tested in a phase 2-3 efficacy trial, can be a major advancement in the prophylaxis and clinical management of COVID-19, because of its broad spectrum, ease of use in non-hospital settings and economic sustainability.

## Introduction

The COVID-19 pandemic highlighted the potential of human monoclonal antibodies (mAbs) to tackle pandemics as they demonstrated to be safe and effective therapeutic tools that can be brought from discovery to proof-of-concept trials in only 5 – 6 months^1^. Since the start of the pandemic a dozen mAbs capable of neutralising the SARS-CoV-2 virus have been identified and are under clinical development. Regulatory agencies have granted Emergency Use Authorization (EUA) to Eli Lilly’s mAb bamlanivimab (LY-CoV555) and mAb combination bamlanivimab + etesevimab, to Regeneron’s mAb combination casiribimab + imdevimab (REGN-COV2), to Celltrion regdanvimab, and GSK/VIR sotrovimab.^2-5^ With the emergence of antibody-resistant SARS-CoV-2 variants of concern (VoCs) at the end of 2020, some mAbs lost their clinical efficacy and in April 2021 the FDA revoked EUA for bamlanivimab.^6^ The Toscana Life Sciences Foundation has recently reported the isolation and characterization of the neutralising mAb, MAD0004J08 from a convalescent COVID-19 patient.^7^ MAD0004J08 appears to be the best candidate for clinical development as it displays the most desirable characteristics for the development of mAb-based prophylaxis and therapy: 1) unprecedented high neutralising potency, implying low dose requirement, intramuscular route of administration, and significant cost reduction of cost of goods. 2) breadth of neutralising activity, *in vitro* vs. the original Wuhan virus and all current SARS-CoV-2 VoC; B.1.1.7 (alpha), B.1.351 (beta), B.1.1.248 (gamma) and B.1.617.2 (delta) with a potency below 10 ng/mL (^7^ and manuscript in preparation), implying broad clinical usefulness in countries with diverse SARS-CoV-2 epidemiology. 3) engineered immunoglobulin fragment crystallizable (Fc) region to increase its serum half-life while silencing the Fc activity to abrogate binding to FcγRs and eliminate possible risks of antibody dependent enhancement (ADE) of disease.^7^

Due to these unique properties, MAD0004J08 is expected to accelerate clearance of the virus, not induce proinflammatory cytokine response, and thus prevent clinical deterioration of COVID-19 disease in patients.

This paper reports the results of the first 30 days of an ongoing phase 1, placebo controlled, double-blind, randomized, single dose, dose-escalation study to evaluate the safety, pharmacokinetics and virus neutralisation titres of the anti-SARS-CoV-2 monoclonal antibody MAD0004J08 in 30 healthy adults. The data generated thus far suggest MAD0004J08 has a strong potential as global therapeutic tool against COVID-19.

## Methods

Details on methods can be found in the study protocol (Addendum).

### Trial design

Our phase 1, first in human (FIH) trial is underway at two sites in Italy (Istituto Nazionale Malattie Infettive Lazzaro Spallanzani, Rome, and Centro Ricerche Cliniche di Verona s.r.l. (CRC), Verona). The final protocol and informed consent were approved by the institutional review boards of each of the participating investigational sites. This study is designed and conducted in accordance with the Declaration of Helsinki, the current revision of Good Clinical Practice (GCP), ICH topic E6 (R2), and the applicable local law requirements. The trial is registered with EudraCT N.: 2020-005469-15 and ClinicalTrials.gov Identifier: NCT04932850.

This is a dose escalation study, open label across doses and randomized, double blind withing each dose level. A total of 30 healthy men and nonpregnant women, 18 to 55 years of age, meeting all inclusion/exclusion criteria were enrolled in three sequential cohorts of 10 subjects each. Within each cohort subjects were randomized with 4:1 ratio to a single i.m. dose of MAD0004J08 (48 mg in Cohort 1, 100 mg in Cohort 2, and 400 mg in Cohort 3) or placebo using an interactive web response system (IWRS). Within each cohort subjects were grouped in two groups of five: the five subjects of the first group, referred to as “sentinels” were randomized one at a time at 48-hour intervals, assuming no safety concern in the investigator’s judgement; the five subjects of the second group were randomized and enrolled with no time restriction. An Independent Data Safety Monitoring Board (DSMB) recommended progress from Cohort 1 to Cohort 2 and from Cohort 2 to Cohort 3 based pre-defined criteria.^8^

Each subject is to undergo 12 visits (V) over a 6-month period: V1-V2 for screening procedures, V1-V3 (Days 1-3) as inpatient in the study center, and V4-V12 as outpatient for follow-up. On day 1 (V3), a single 5mL injection was administered in the right gluteus for cohorts 1 and 2 and two 5mL injections, were administered one in the right and one left gluteus for cohort 3. Each subject was provided with two diaries to self-report and record solicited adverse events (AEs) from day 1 to day 7, and unsolicited AEs concomitant medication throughout the study

### Eligibility Criteria

The detailed list of inclusion/exclusion criteria can be found in the trial-protocol. Key inclusion criteria were as follows: age 18-55, signed informed consent, willingness to use appropriate contraception, body mass index-18·5-30 kg/m2; systolic blood pressure 90-139 mmHg, diastolic blood pressure 69-90 mmHg; heart rate 50-100 bpm; electrocardiogram (ECG) without clinically significant abnormalities; negative SARS-CoV-2 serology test (negative anti-S and anti-N) or negative SARS-CoV-2 qRT-PCR in the 72h prior with result before the treatment. Key exclusion criteria were as follows: prior intake of investigational or licensed vaccine for the prevention of SARS CoV2; history of infection with SARS or MERS; positive or missing pregnancy test at screening or day 1 or lactating women; history of allergic reactions likely to be exacerbated by any component of the investigational product; previous intake of a mAb within 6 months; history of malignancy in the last 5-7 years; Immunodeficiency due to illness, any course of glucocorticoid therapy exceeding 2 weeks; acute illnesses-history of renal, hepatic, gastrointestinal, cardiovascular, respiratory, dermatologic, hematological, endocrine, psychiatric or neurological diseases that may interfere with the aim of the study or increase subjects risks in investigator’s opinion. A statement was included in the study protocol that if during the study, a subject is included in a vaccination list according to the national guidelines, the best option for the subject will be pursued. However, such situation did not arise during the study.

### Investigational medicinal product (IMP)

MAD0004J08, human monoclonal antibody 100 mg / 2·5 mL solution for injection was manufactured by Menarini Biotech, and was characterized and filled, at Istituto Biochimico Italiano (IBI) Lorenzini. The placebo was prepared as a 2·5mL of a 0·9% w/v sterile solution of sodium chloride (NaCl) in water for injections (WFI) by IBI. The investigational products were to be stored at 2-8°C in a dry locked place.

### End Points

In this report, results from the following study end points are presented. The full set of endpoints (see protocol) in the complete sample will be presented upon completion of the study.

Primary end point: Proportion of subjects with severe and/or serious treatment-emergent adverse events (TEAEs), including clinically relevant laboratory abnormalities, vital signs, and adverse reactions at the injection site) in the 7 days post-treatment. A TEAE is defined as any AE with onset after administration of study drug (N=30).

Secondary end points: Proportion of subjects with solicited local AEs (pain, redness and swelling at injection site) and systemic AEs (headache, fatigue, muscle pain, joint pain, vomiting, diarrhea, chills, fever) from day 1 to day 7 (N=30); MAD0004J08 sera concentrations on day 1 (0, 1, 2, 3, 4, 6, 8, 12, and 24h), days 2, 8, 15, 22, 30 (N=24; subjects considered placebo were excluded from the analyses); MAD0004J08 sera neutralising ability at baseline, and on days 2, 8, and 30 (N=24; subjects considered placebo were excluded from the analyses).

### ELISA for PK analyses

The method for detection of MAD0004J08 in human serum is a quantitative sandwich ELISA. Briefly, a 96-wells plate is coated with SARS-CoV-2 Spike protein; after blocking of the plate with PBS + 1% BSA, samples containing MAD0004J08 are pipetted into the wells, followed by a wash with PBS + 0.05% Tween-20 to remove all unbound matrix components; alkaline phosphatase labelled anti-Human IgG (γ chain specific) is added to bind to the immobilized MAD0004J08; the complex is detected by pNPP substrate; after a wash to remove unbound reagents, the enzyme is revealed by its action on the pNPP substrate; after stopping the reaction with a strong base, the intensity of the color (read at 405 nm) is directly proportional to the amount of MAD0004J08 present in the sample.

### SARS-CoV-2 viruses

The SARS-CoV-2 viruses used to perform the neutralisation assay were SARS-CoV-2 wild type (EVAg GenBank: MT066156.1), SARS-CoV-2 B.1.1.7 (INMI GISAID accession number: EPI_ISL_736997), SARS-CoV-2 B.1.351 (EVAg Cod: 014V-04058) and B.1.1.248 (EVAg CoD: 014V-04089).

### SARS-CoV-2 neutralisation assay

SARS-CoV-2 neutralisation assay was performed as previously described.^7^ Briefly, plasma samples were tested at a starting dilution of 1:10 and then diluted in steps of 1:2 for twelve points. All samples were mixed with a SARS-CoV-2 wild type, SARS-CoV-2 B.1.17 (alpha), B.1.351 (beta) or B.1.1.248 (gamma) viral solution containing 100 TCID_50_ of the virus. After 1 hour incubation at 37°C, 5% CO_2_, virus-mAb mixture was added to the wells of a 96-well plate containing a sub-confluent Vero E6 cell monolayer. Plates were incubated for 3 - 4 days at 37°C in a humidified environment with 5% CO_2_, then examined for CPE by means of an inverted optical microscope. Technical duplicates were performed for each experiment.

Neutralising activities are represented as geometric mean neutralisation titres (GMT) leading to hundred percent viral neutralisation/inhibition (ID_100_). In the original protocol only GMT against the wild type (WT) Wuhan stain were envisaged. As post-hoc analyses, in light of the evolving epidemiology GMTs vs. alpha, beta and gamma variants of concern were also assessed. In addition, as post hoc analyses, GMTs following administration of MAD0004J08 were compared to matching GMTs obtained from pools of convalescent and vaccinated sera.

### Plasma specimens from COVID-19 convalescent and vaccinated subjects

Plasma specimens from COVID-19 convalescent subjects and SARS-COV-2 vaccinated subjects used as comparators in the present paper were previously collected from a separate study conducted in collaboration with the National Institute for Infectious Diseases, IRCCS – Lazzaro Spallanzani Rome (IT) and Azienda Ospedaliera Universitaria Senese, Siena (IT). The study was approved by local ethics committees and conducted according to good clinical practice Plasma neutralisation titres of COVID-19 convalescent subjects were previously reported by Andreano et al.^9^

### Statistical Analysis

To assess the pharmacokinetics of MAD0004J08 in the three different cohorts, a non-parametric Mann–Whitney T test was performed among groups at all different time points. For the analyses of sera or plasma neutralisation antibody titres against SARS-CoV-2 and VoCs, the 100% inhibitory dilution (ID_100_) was calculated as geometric mean of two technical duplicates and statistical significances of the differences among groups was determined by non-parametric Mann–Whitney T test using GraphPad Prism software (version 8.0.2). Significances were denoted in each figure and shown as * (p≤0·05), ** (p≤0·01), *** (p≤0·001) and **** (p≤0·0001).

## Results

Trial was initiated by screening 50 healthy volunteers between March and May 2021. There were a total of 15 screen failures, 13 due to inclusion/exclusion criteria not met and two to withdrawal. Five screened volunteers were kept as reserves. Thirty were enrolled and randomised. Baseline demographic characteristics of the participants in the safety population at enrolment were similar among the treatment groups in terms of sex, mean age, and race (Table 1). Overall, the study population consisted of white healthy male and female participants with a mean age of 32·2 years (range 19–54 years); 56·6% were male and 43·4% were female (Table 1).

**Table 1:**
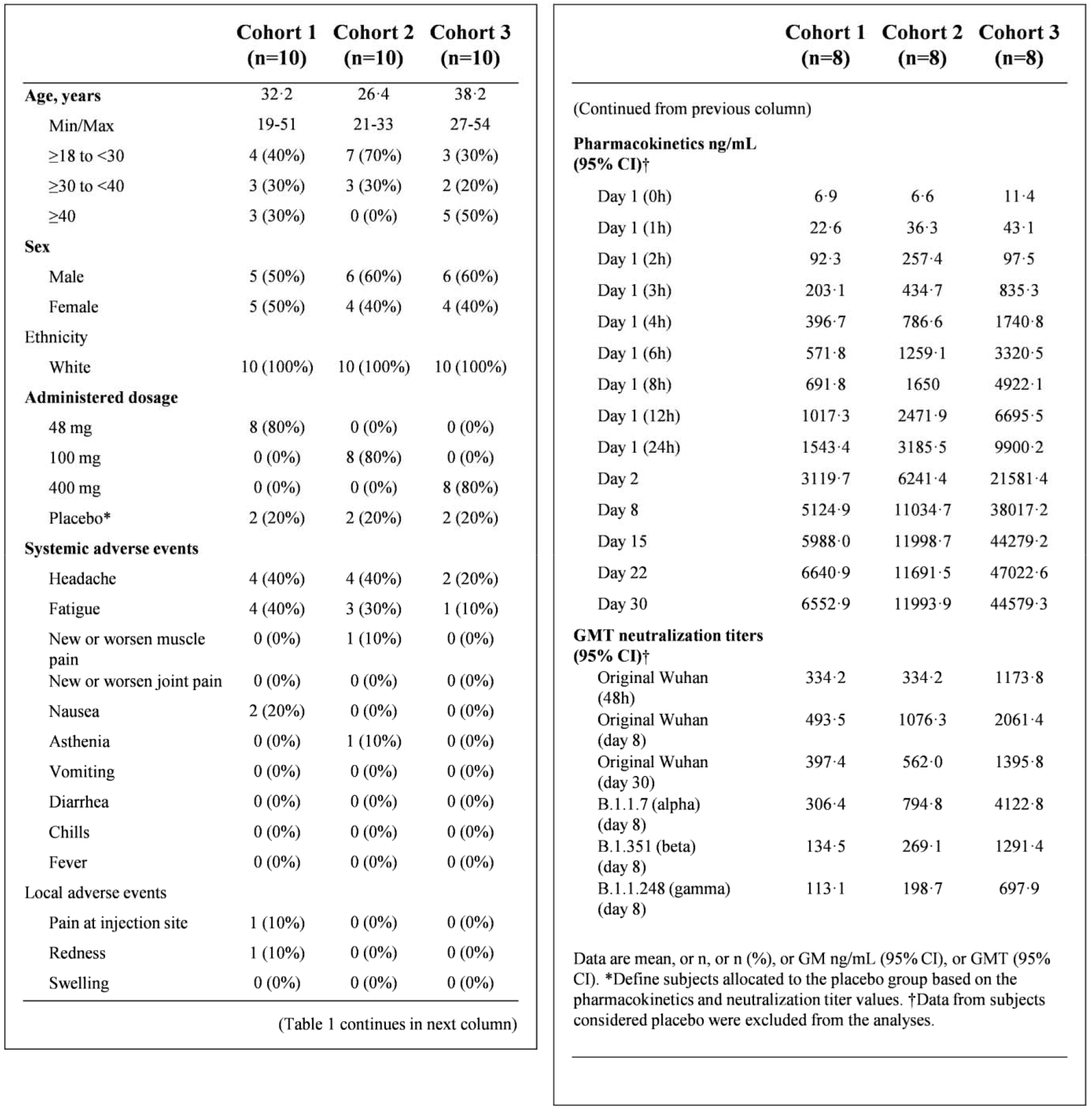
MAD0004J08 phase I clinical trial summary.

|  | Cohort 1<br>(n=10) | Cohort 2<br>(n=10) | Cohort 3<br>(n=10) |
| --- | --- | --- | --- |
| **Age, years** | 32·2 | 26·4 | 38·2 |
| Min/Max | 19-51 | 21-33 | 27-54 |
| ≥18 to <30 | 4 (40%) | 7 (70%) | 3 (30%) |
| ≥30 to <40 | 3 (30%) | 3 (30%) | 2 (20%) |
| ≥40 | 3 (30%) | 0 (0%) | 5 (50%) |
| **Sex** |  |  |  |
| Male | 5 (50%) | 6 (60%) | 6 (60%) |
| Female | 5 (50%) | 4 (40%) | 4 (40%) |
| **Ethnicity** |  |  |  |
| White | 10 (100%) | 10 (100%) | 10 (100%) |
| **Administered dosage** |  |  |  |
| 48 mg | 8 (80%) | 0 (0%) | 0 (0%) |
| 100 mg | 0 (0%) | 8 (80%) | 0 (0%) |
| 400 mg | 0 (0%) | 0 (0%) | 8 (80%) |
| Placebo* | 2 (20%) | 2 (20%) | 2 (20%) |
| **Systemic adverse events** |  |  |  |
| Headache | 4 (40%) | 4 (40%) | 2 (20%) |
| Fatigue | 4 (40%) | 3 (30%) | 1 (10%) |
| New or worsen muscle pain | 0 (0%) | 1 (10%) | 0 (0%) |
| New or worsen joint pain | 0 (0%) | 0 (0%) | 0 (0%) |
| Nausea | 2 (20%) | 0 (0%) | 0 (0%) |
| Asthenia | 0 (0%) | 1 (10%) | 0 (0%) |
| Vomiting | 0 (0%) | 0 (0%) | 0 (0%) |
| Diarrhea | 0 (0%) | 0 (0%) | 0 (0%) |
| Chills | 0 (0%) | 0 (0%) | 0 (0%) |
| Fever | 0 (0%) | 0 (0%) | 0 (0%) |
| **Local adverse events** |  |  |  |
| Pain at injection site | 1 (10%) | 0 (0%) | 0 (0%) |
| Redness | 1 (10%) | 0 (0%) | 0 (0%) |
| Swelling | 0 (0%) | 0 (0%) | 0 (0%) |

|  | Cohort 1<br>(n=8) | Cohort 2<br>(n=8) | Cohort 3<br>(n=8) |
| --- | --- | --- | --- |
| (Continued from previous column) |  |  |  |
| **Pharmacokinetics ng/mL (95% CI)†** |  |  |  |
| Day 1 (0h) | 6·9 | 6·6 | 11·4 |
| Day 1 (1h) | 22·6 | 36·3 | 43·1 |
| Day 1 (2h) | 92·3 | 257·4 | 97·5 |
| Day 1 (3h) | 203·1 | 434·7 | 835·3 |
| Day 1 (4h) | 396·7 | 786·6 | 1740·8 |
| Day 1 (6h) | 571·8 | 1259·1 | 3320·5 |
| Day 1 (8h) | 691·8 | 1650 | 4922·1 |
| Day 1 (12h) | 1017·3 | 2471·9 | 6695·5 |
| Day 1 (24h) | 1543·4 | 3185·5 | 9900·2 |
| Day 2 | 3119·7 | 6241·4 | 21581·4 |
| Day 8 | 5124·9 | 11034·7 | 38017·2 |
| Day 15 | 5988·0 | 11998·7 | 44279·2 |
| Day 22 | 6640·9 | 11691·5 | 47022·6 |
| Day 30 | 6552·9 | 11993·9 | 44579·3 |
| **GMT neutralization titers (95% CI)†** |  |  |  |
| Original Wuhan (48h) | 334·2 | 334·2 | 1173·8 |
| Original Wuhan (day 8) | 493·5 | 1076·3 | 2061·4 |
| Original Wuhan (day 30) | 397·4 | 562·0 | 1395·8 |
| B.1.1.7 (alpha) (day 8) | 306·4 | 794·8 | 4122·8 |
| B.1.351 (beta) (day 8) | 134·5 | 269·1 | 1291·4 |
| B.1.1.248 (gamma) (day 8) | 113·1 | 198·7 | 697·9 |
| Data are mean, or n, or n (%), or GM ng/mL (95% CI), or GMT (95% CI). *Define subjects allocated to the placebo group based on the pharmacokinetics and neutralization titer values. †Data from subjects considered placebo were excluded from the analyses. |  |  |  |
| **Table 1: MAD0004J08 phase I clinical trial summary** |  |  |  |

No severe or serious treatment emergent adverse event (TEAE) was reported through 7 days post dosing. Local and systemic solicited adverse events through day 7 occurred in few subjects (Table 1), were all mild to moderate, lasted no more than 2 and 6 days for systemic and local TEAE respectively, and showed no sign of dose-related increase of frequency or severity. Overall, systemic solicited adverse events (n=22; 55·0%) were more frequent than local solicited adverse events (n=2; 6·7%) (Table 1).

The baseline (pre-dose) geometric mean (GM) concentrations at day 1 (0h) of anti-spike antibodies measured by ELISA were at background levels. Two subjects/cohort had GM levels below the lower limit of quantification (LLOQ) for all samples. They were assumed to belong to the placebo group and were not considered in the analysis. Detectable levels of antibodies were seen as quickly as two hours post-administration and increased constantly in a dose-dependent manner. At day 8, mean sera binding titres almost peaked showing a GM of 5124·9, 11034·7, and 38017·2 ng/mL for cohorts 1, 2, and 3 respectively. Sera binding titres continued to increase up to 30 days with a geometric mean of 6552·9, 11993·9, 44579·3 ng/mL for cohorts 1, 2, and 3 respectively (Figure 1; Table 1 and Supplementary Table 1).

**Figure 1:**
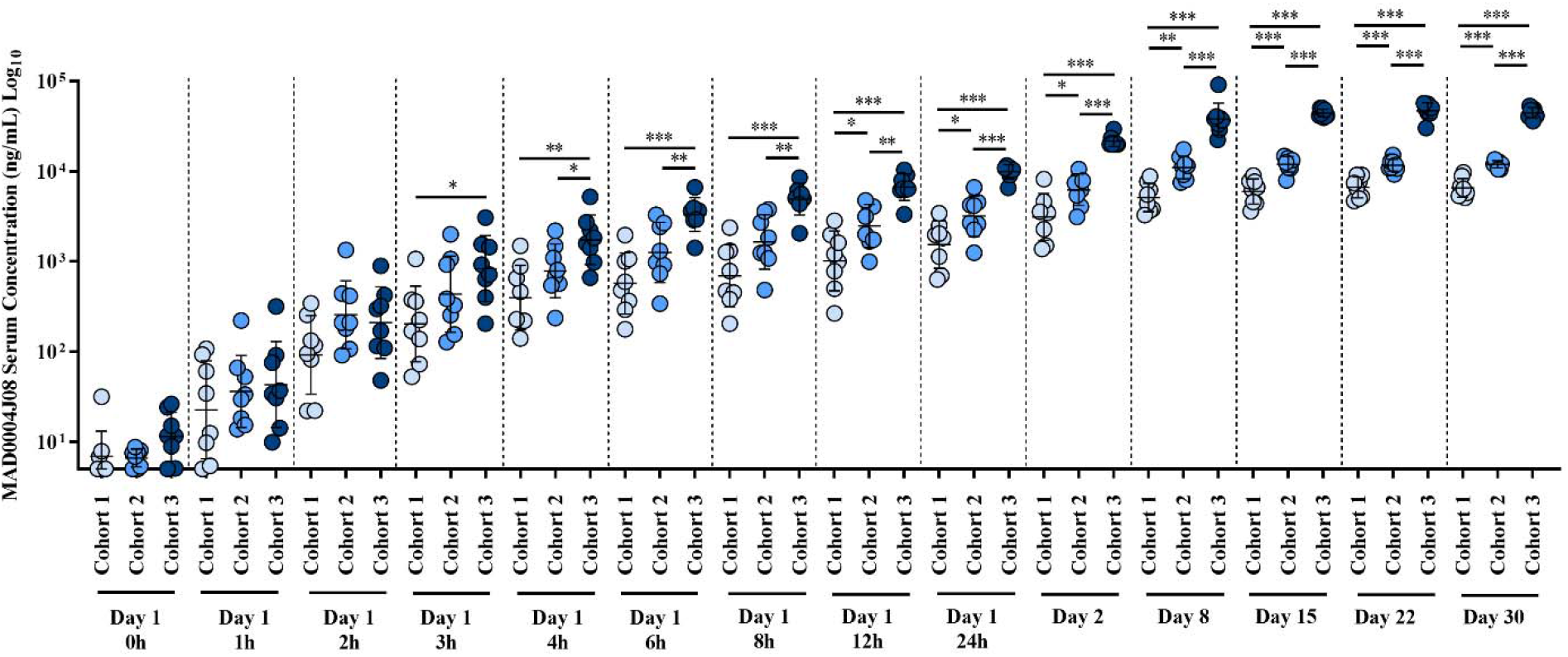
MAD0004J08 pharmacokinetics. SARS-CoV-2 spike (S) protein IgG mean titers in (ng/mL) pre and post MAD0004J08 injection in three cohorts. Spots indicated the individual antibody titers, Cohort 1 (48mg-light blue), Cohort 2 (100mg-blue) and Cohort 3 (400mg-dark blue). Only p values for statistically significant differences are shown in the figure. Under the blinded condition, 2 subjects / cohort with very low/ undetectable levels of serum titers against SARS-Cov-2-S protein have been assumed to belong to the placebo arm and are not shown in the figure.

The study assessed the neutralising activity of sera from MAD0004J08 immunized subjects against SARS-CoV-2 original Wuhan virus. The results are represented as geometric mean neutralisation titres (GMT) leading to hundred percent viral neutralisation/inhibition (ID_100_). The geometric mean neutralisation titres (GMT) in MAD0004J08 administered subjects at baseline were below the lower limit of quantitation (LLOQ-less than 10). Two subjects/cohort had GMT levels below the LLOQ at all subsequent time points and were assumed to belong to the placebo group and were not considered in the analysis. The GMT levels in cohort 1 (48mg) on day 2 was 334·2, increased to 493·5 on day 8 and was 397·4 on day 30. The GMT in cohort 2 (100mg) was 334·2 on day 2, improved further to 1076·3 on day 8 and was 562·0 on day 30. The GMT levels for cohort 3 (400mg) were 1173·8, 2061·4 and 1395·8 on day 2, 8 and 30 respectively (Table 1; Supplementary Table 2).

Additionally, we compared the neutralising activity of sera from MAD0004J08 immunized subjects with COVID-19 convalescent patients (n=20) and two groups of BNT162b2 mRNA vaccine recipients (n=5/group) who were either seronegative or seropositive for SAR-CoV-2 before vaccination. The GMT for 20 COVID-19 convalescent patients had a wide GMT range 1-10,240 which can be explained by differences in clinical severity of COVID-19 disease as 15/20 (75%) were hospitalized and 10/15 (66·7%) of hospitalized patients were on oxygen therapy. The neutralisation GMT for all 20 COVID-19 convalescent patients was 73·7, whereas the average GMT for seronegative and seropositive vaccinees were 27·0 and 269·3, respectively (Figure 2; Supplementary Table 3). Overall, the neutralising ability of sera from cohort 1, 2 and 3 subjects at day 8 were 6·7-, 14·6-, 27·9-fold higher than COVID 19 convalescent sera and were 18·3-, 40·0-,76·4- and 1·83-, 4·0-, 7·6-fold significantly higher compared to seronegative and seropositive vaccinees respectively (Figure 2; Supplementary Table 3).

**Figure 2:**
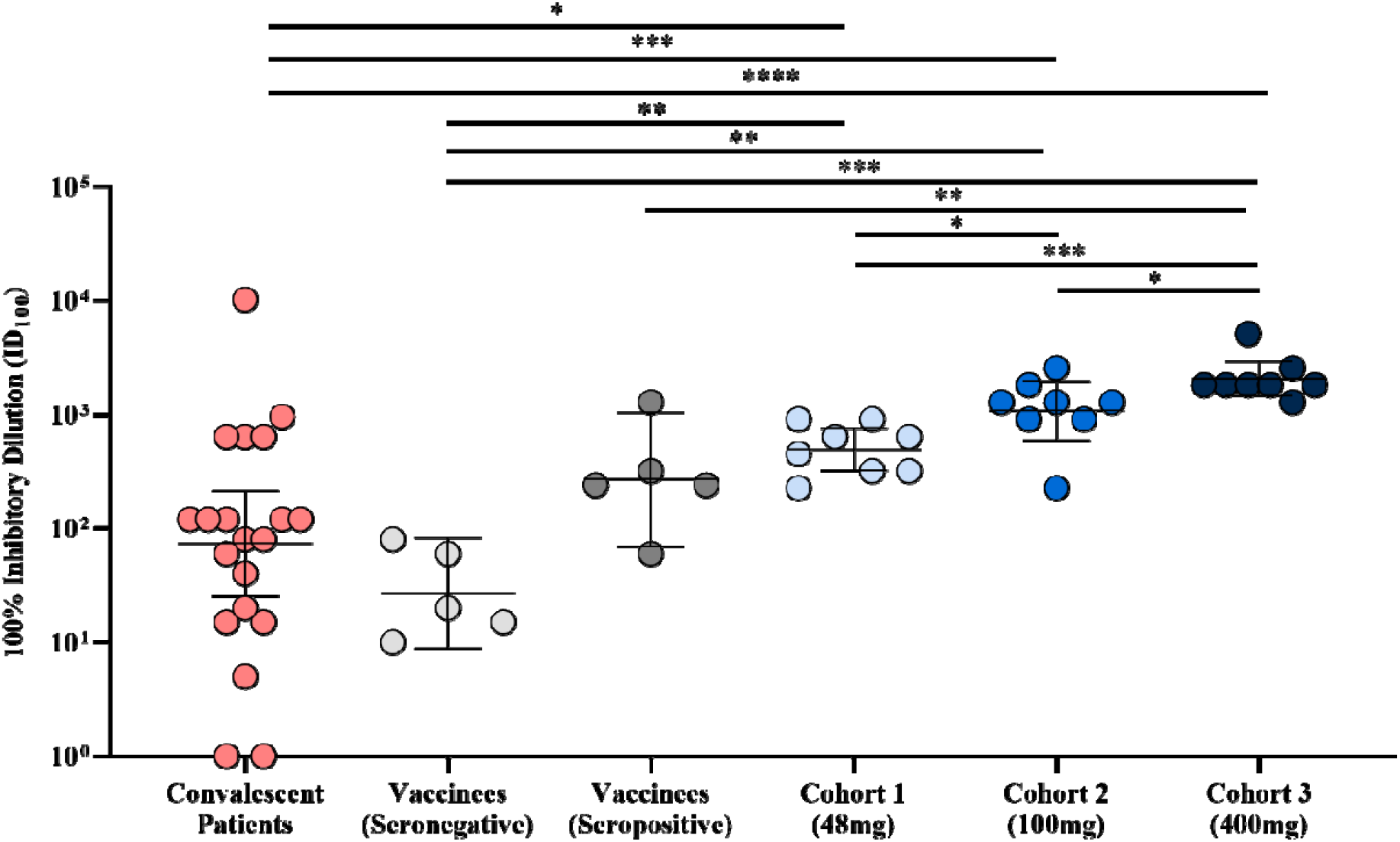
Serum neutralisation activity against SARS-CoV-2 Wuhan virus. The graph shows the neutralisation reported as 100% inhibitory dilution (ID_100_) of sera collected from COVID-19 convalescent patients, vaccinees not exposed (seronegative) or previously exposed (seropositive) to SARS-CoV-2 and subjects that received MAD0004J08 at 48, 100 and 400 mg. Significances are shown as * (p≤0.05), ** (p≤0.01), *** (p≤0.001) and **** (p≤0.0001).

Finally, we evaluated the sera neutralisation activity on day 8 of all subjects that received MAD0004J08 against the SARS-CoV-2 variants of concern B.1.1.7 (alpha), B.1.351 (beta) and B.1.1.248 (gamma) (Figure 3). GMT and 95% confidence interval (95% CI) are reported in Table 1 and Supplementary Table 2. Overall, a single *i.m.* injection of MAD0004J08 resulted in high neutralization titres against all tested SARS-CoV-2 variants in a dose-dependent fashion with GMT levels for cohort 1 of 306·4, 134·5 and 113·1, for cohort 2 of 794·8, 269·1 and 198·7, and for cohort 3 of 4122·8, 1291·4 and 697·9 against the B.1.1.7, B.1.351 and B.1.1.248 respectively (Figure 3A - C; Table 1; Supplementary Table 2). GMT levels against the B.1.1.7 variant were comparable to the SARS-CoV-2 Wuhan virus for cohort 1 and 2, while the GMT was 2-fold higher for cohort 3 (Figure 3D - F). Neutralisation titres compared to the original Wuhan virus dropped by 3·67-, 4·00- and 1·68-fold against B.1.351 (beta) and by 4·36-, 5·42- and 2·95-fold against B.1.1.248 (gamma) for cohort 1, 2 and 3 respectively (Figure 3D – F; Table 1; Supplementary Table 2). However, despite reductions in neutralisation titres were observed against the variants, GMT levels were either comparable in case of cohort 1 (48mg) or significantly higher for cohort 2 and 3 (100 and 400mg) than those observed in convalescent patients and vaccinees against the Wuhan virus (Figure 2 - 3).

**Figure 3:**
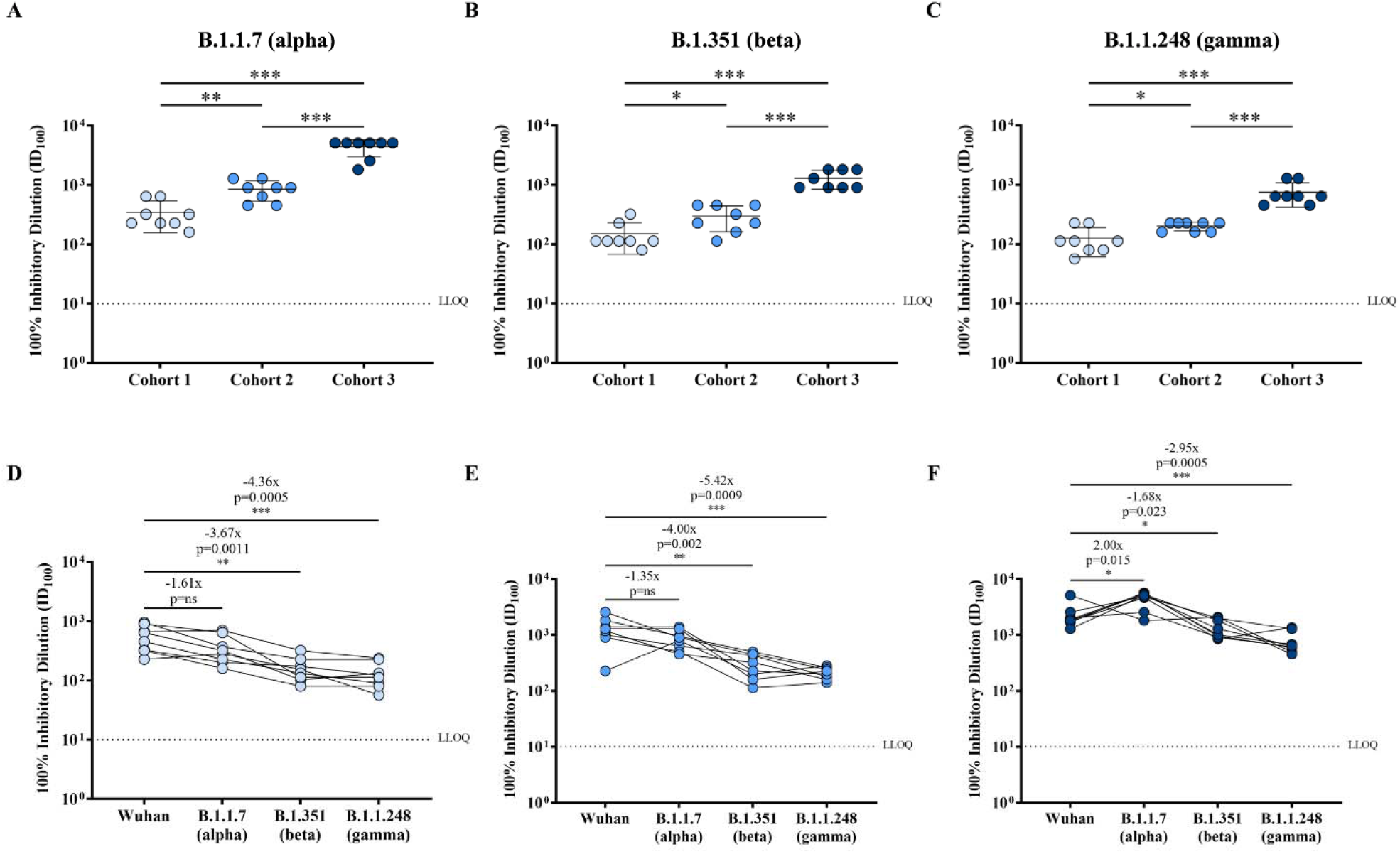
Serum neutralisation activity against SARS-CoV-2 variants of concern. (A – C) Graphs show the neutralisation activity reported as 100% inhibitory dilution (ID_100_) of sera collected from subjects who received MAD0004J08 at 48, 100 and 400 mg against SARS-CoV-2 B.1.1.7 (A), B.1.351 (B) and B.1.1.248 (C) variants. (D – F) Graphs show fold changes of serum neutralisation activity against the different SARS-CoV-2 VoCs tested in this study for Cohort 1 (D), Cohort 2 (E) and Cohort 3 (F). On each graph serum neutralisation titre fold changes against VoCs are reported with respect to the Wuhan virus. Significances are shown as * (p≤0.05), ** (p≤0.01), *** (p≤0.001) and **** (p≤0.0001).

## Discussion

The results of this study show that, following a single *i.m.* administration, MAD0004J08 at 48, 100 and 400mg is safe, well-tolerated with minimal reactogenic adverse events, and rapidly distributes systemically. In addition, a single dose of MAD0004J08 resulted in extremely high serum neutralization titers which were 5- to 76-fold higher compared to COVID-19 convalescent donors and BNT162b2 mRNA vaccinees. With the emergence and worldwide spread of SARS-CoV-2 variants of concern it is imperative to develop mAbs able to neutralise circulating variants of concern (VoC), therefore maintaining their therapeutic efficacy. Some mAbs already in clinical application for the treatment of COVID-19, showed to fall short against emerging variants and for one of them due to lack of clinical efficacy regulatory agencies revoked emergency use authorization.^6^ In this study we showed that sera from MAD0004J08 treated subjects retained high neutralization titres against major SARS-CoV-2 VoCs including the B.1.1.7 (alpha), B.1.351 (beta) and B.1.1.248 (gamma) variants highlighting the potential of this antibody to be used globally for the treatment of COVID-19. Although sera from 400mg dose showed the highest neutralisation titres against SARS-CoV-2 and variants of concern, the 100 mg dose, which was also found to have significantly higher neutralising titres compared to convalescent patients and vaccinees, could be sufficient and strategically more suitable to develop. In fact, a single 100mg dose of MAD0004J08 could increase the number of available doses to give to COVID-19 patients.

To complement the ability of MAD0004J08 to neutralise major SARS-CoV-2 VoC the Fc modifications introduced to extend its serum half-life and to limit risk of ADE in hospitalized COIVD-19 patients, could further increase the clinical suitability of this antibody. Concerning the serum half-life extension, our data showed that sera neutralization titers were maintained at high levels up to the 30 days post-administration and follow-up is planned up to 6 months. As for the silencing of the Fc-portion to minimize risks of ADE our strategy, compared to the use of unmodified or enhanced Fc-functions in antibodies already in clinic, could result to be successful in hospitalized COVID-19 patients with and without high titres of serum antibodies against SARS-CoV-2. In fact, at the end of 2020, the Eli Lilly bamlanivimab (LY-CoV555) received emergency used authorization but application was limited as the administration of this antibody may be associated with worse clinical outcomes when administered to hospitalized patients requiring high flow oxygen or mechanical ventilation with COVID-19.^5^ In addition, a recent primary clinical trial under peer-review has shown that intravenous administration of REGEN-COV mAb cocktail, at a total dose of 8g in hospitalized COVID-19 patients reduced 28-day mortality among patients who were seronegative at baseline, highlighting important role of mAbs therapy even in hospitalized COVID-19 patients.^10^ However, COVID-19 seropositive hospitalized patients did not benefit from REGEN-COV administration, suggesting that presence of high antibody titres induced by infection plus the infusion of enormous quantity of mAbs for therapy may not be beneficial for these patients and ADE mechanisms could be involved. A recent paper describing the longitudinal neutralising antibodies in patients progressing to severe disease, indicates antibody dependent enhancement as one of the mechanisms of increased disease severity.^11^ These previous reports indicate that removing Fc-functions could result to be a successful strategy for the treatment of hospitalized seronegative as well as seropositive COVID-19 patients.

The limitation of our study is that our first in human study was conducted in healthy population of age 18-55 without much diversity, whereas the risk for severity of COVID-19 disease is more in population with comorbid conditions or older age groups. Our ongoing Phase 2-3 study will assess MAD0004J08 in mild/moderate diseased and stratified subjects groups to assess dose selection and efficacy.

To conclude, MAD0004J08 administration is safe, confers broad-coverage against major SARS-CoV-2 variants of concern, and giving the low dosage needed and *i.m.* route of administration can be a globally available and affordable countermeasure to the COVID-19 pandemic.

## Data Availability

All metadata generated in this analysis will be available.

## Contributors

SL, GDC, GI and RR conceptualised the study. SL, SM, AA, EN, EG, MMP, EA, IP, GP, AG, IH and ML contributed to acquisition of data. EA, AP, AG, SN and RR contributed to data analysis and interpretation. EA, AP and RR drafted the manuscript. All authors revised the manuscript, read and approved the final version. AP, AF and RR acquired funding for the study. SN, AT, GP, EM, GI, FV and RR contributed to the logistic of the study. All authors of the manuscript had access to all the data in the study.

## Data Sharing

All metadata generated in this analysis will be available.

## Acknowledgments

The authors would like to thank the donors, physicians, and staff participating in this phase I clinical trial aimed to evaluate safety and tolerability of MAD0004J08. We would also like to thank CROss Allience, Mendrisio, Svizzera, Ardena Bioanalyses, Assen, Netherlands, ExcellGene, Monthey, Switzerland, Menarini Biotech S.r.l., Pomezia, Italy and Istituto Biochimico Italiano (IBI) Lorenzini, Aprilia, Italy, for their support to this study. The authors would also like to acknowledge Concetta Castelletti (laboratory), clinical team: Ilaria Mastrorosa, Alessandra Vergori, Sandrine Ottou, Serena Vita, Laura Scorzolini, Alessandra D’Abramo.

This study was supported by the EU Malaria Fund, inaugurated on the 3^rd^ of June 2020 and initiated by the kENUP Foundation. This publication was supported by the COVID-2020-12371817 project, which has received funding from the Italian Ministry of Health (Ministero della Salute), from the Italian Ministry of Economic (Ministero dello Sviluppo Economico) through the “Contratti di Sviluppo” funding program, and from Regione Toscana. This work was funded by the European Research Council (ERC) advanced grant agreement number 787552 (vAMRes). This publication was supported by the European Virus Archive goes Global (EVAg) project, which has received funding from the European Union’s Horizon 2020 research and innovation program under grant agreement No 653316.

## SUPPLEMENTARY TABLES

**Supplementary Table 1:**
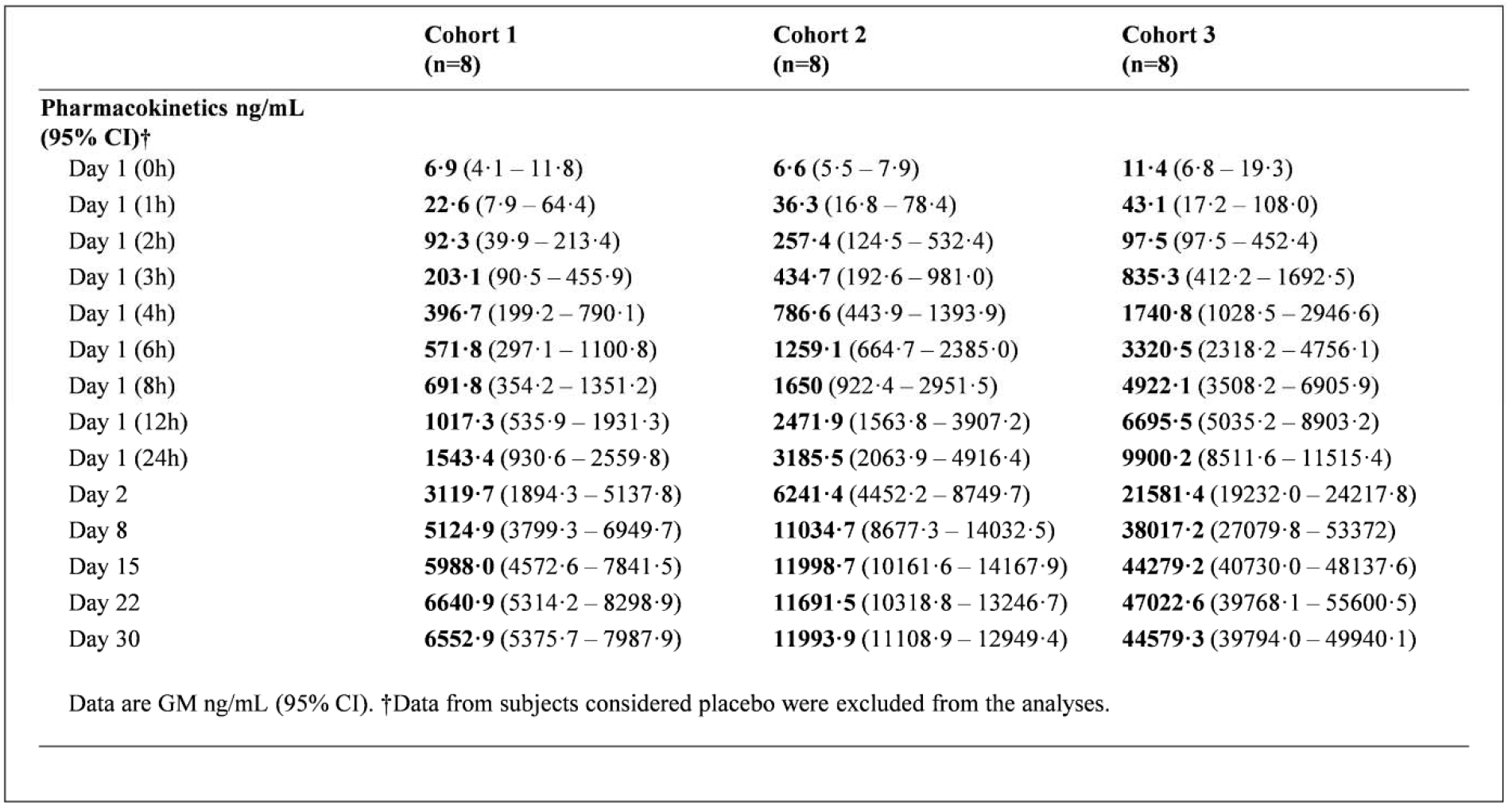
MAD0004J08 pharmacokinetics.

**Supplementary Table 2:**
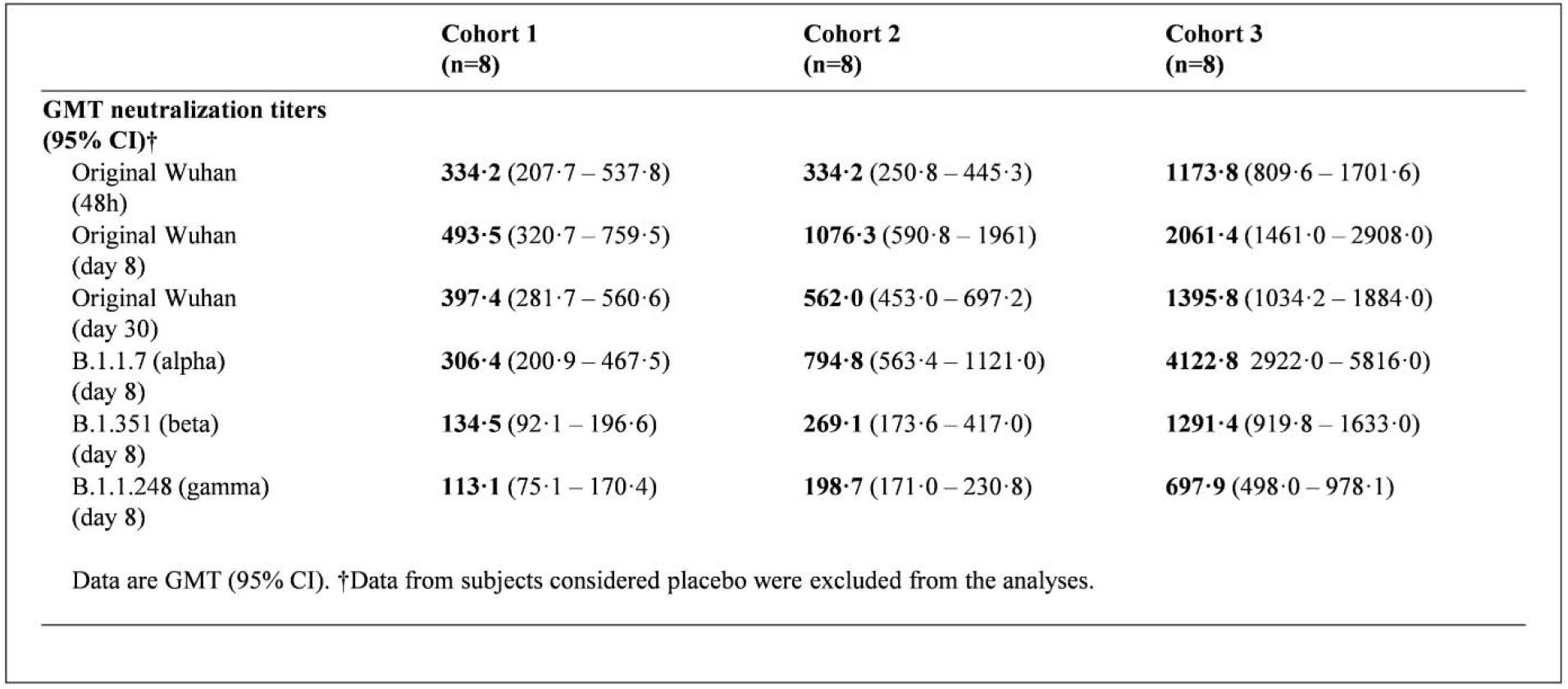
MAD0004J08 serum neutralisation titres.

**Supplementary Table 3:**
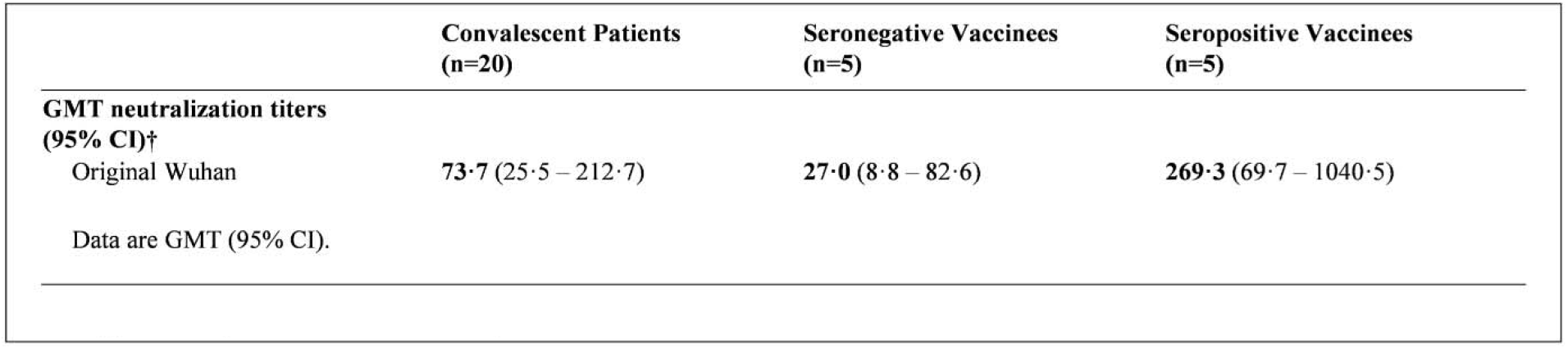
Convalescent patients and COVID-19 BNT162b2 mRNA vaccinees serum neutralization titres.

|  | Convalescent Patients<br>(n=20) | Seronegative Vaccinees<br>(n=5) | Seropositive Vaccinees<br>(n=5) |
| --- | --- | --- | --- |
| **GMT neutralization titers**<br>**(95% CI)†** |  |  |  |
| Original Wuhan | 73·7 (25·5 – 212·7) | 27·0 (8·8 – 82·6) | 269·3 (69·7 – 1040·5) |
| Data are GMT (95% CI). |  |  |  |

